# Visible Immigration Enforcement Arrests and Mental Health Among Hispanic Adults in the United States

**DOI:** 10.64898/2026.07.02.26357125

**Authors:** Erkmen G. Aslim, Dinara Tekin, Atheendar S. Venkataramani

## Abstract

Visible immigration enforcement in community settings can create fear and distress that spill beyond the communities directly affected. In this paper we explore the relationship between state-level community-based U.S. Immigration and Customs Enforcement (ICE) arrest rates and mental health outcomes among Hispanic and non-Hispanic adults in the United States. We explore this question for two periods, 2023-2024 and 2025, over which visible immigration enforcement moved from a localized to a national phenomenon. In 2023-2024, poor mental health rises as local ICE arrest rates increase. Each additional non-prison arrest per 10,000 non-citizens at the state-level is associated with 0.15 additional poor mental health days per month among Hispanic adults, and with a 0.9 percentage point higher likelihood of reporting any poor mental health days, a 2.2 percent increase relative to the mean, driven mainly by female and lower-educated Hispanic adults. The relationship is null for non-Hispanic adults. By contrast, within-state deviations in ICE arrests are no longer correlated with mental health beyond the national trend in 2025. We posit that this may because ICE activity was more frequent and more synchronized and mental health deteriorated nationally relative in 2025 relative 2023-2024, potentially making local ICE activity less salient or impacts of such activity harder to detect.

## Introduction

In recent years, Immigration and Customs Enforcement (ICE) arrests and detention have increased substantially in the United States, with the detained population rising by nearly 75 percent in 2025 to exceed 68,000 individuals held on any given day.^1^ Beyond the scale of this expansion, the nature of enforcement has shifted in important ways. Historically, interior ICE activity largely consisted of custodial transfers, in which noncitizens were moved to immigration detention following arrest by local or state law enforcement. In contrast, enforcement over the last five years has shifted toward direct apprehensions in neighborhoods and workplaces. Such arrests have increased dramatically, and an increasing proportion of those detained have no prior criminal convictions.^2^ Immigration enforcement is therefore increasingly experienced not primarily within jails, but through visible encounters in neighborhoods, workplaces, and other public or community-facing settings, including hospitals and clinics.^3^

The shift in enforcement strategy has implications that extend beyond those who are directly detained. Arrests in neighborhoods, workplaces, and other everyday settings are more visible and less predictable than custodial transfers from jails, generating fear, uncertainty, and hypervigilance even among communities not directly targeted. Because chronic stress is closely associated with depression, anxiety, and substance use, changes in the scale and composition of immigration enforcement may carry meaningful public health consequences.^4–7^ Beyond these psychological spillovers, immigration enforcement near health care settings may also deter care-seeking, as individuals who fear detention avoid or delay contact with the health system.^8,9^ As enforcement becomes a more prominent feature of daily life in many parts of the United States, understanding its effects on both psychological well-being and access to care is essential for evaluating the broader social costs of contemporary policy.

A growing literature links immigration enforcement to mental health. Qualitative and case studies of worksite raids and parental detention document acute psychological distress among children and families directly exposed to enforcement actions.^10,11^ Quantitative work extends this to broader populations: state-level analyses link restrictive immigration policy environments to worse mental health among Latinos,^14^ comparisons around national policy changes show elevated distress among Hispanic adults,^15^ and survey evidence shows that US citizens with social ties to detained or deported migrants, especially US-citizen Latinos, report more anxiety, depression, and distress.^13^ Additionally, studies of local enforcement document broader health and social disruption in Latino communities.^12,16^ This work, however, largely predates the recent enforcement era, emphasizes policy adoption or isolated raids rather than sustained arrest intensity, and rarely separates community-based arrests from custodial transfers.^17–19^ We build on it using recent, nationally representative data and high-frequency variation in community-based arrests, isolating the most visible form of enforcement now central to ICE activity and identifying the groups whose health is most affected.

The goal of this paper is to provide new evidence on the mental health consequences of contemporary community-based ICE activity. Specifically, we examined whether higher levels of community-level ICE arrests were associated with worse adult mental health outcomes in the United States using recent nationally representative data. We linked individual-level data from the 2023 and 2024 Behavioral Risk Factor Surveillance System (BRFSS) to monthly state-level ICE arrest records from the Deportation Data Project to explore how contemporaneous variation in enforcement intensity is associated with mental health outcomes. We also used 2025 BRFSS data to examine how this relationship changed as enforcement escalated and shifted from a more localized to a national phenomenon. In both periods, we distinguish community-based arrests from custodial transfers to isolate the form of enforcement that has become increasingly central to contemporary ICE activity.

## Results

Table 1 reports mean mental health outcomes for Hispanic and non-Hispanic adult respondents in the BRFSS in states with below-versus above-median ICE arrest rates, separately for 2023-2024 and 2025. In general, Hispanic adults in high-enforcement states reported worse outcomes on most mental health measures compared to Hispanic adults in low-enforcement states (with similar differences seen for some mental health outcomes for non-Hispanic adults). However, standardized differences were generally small.

**Table 1.**
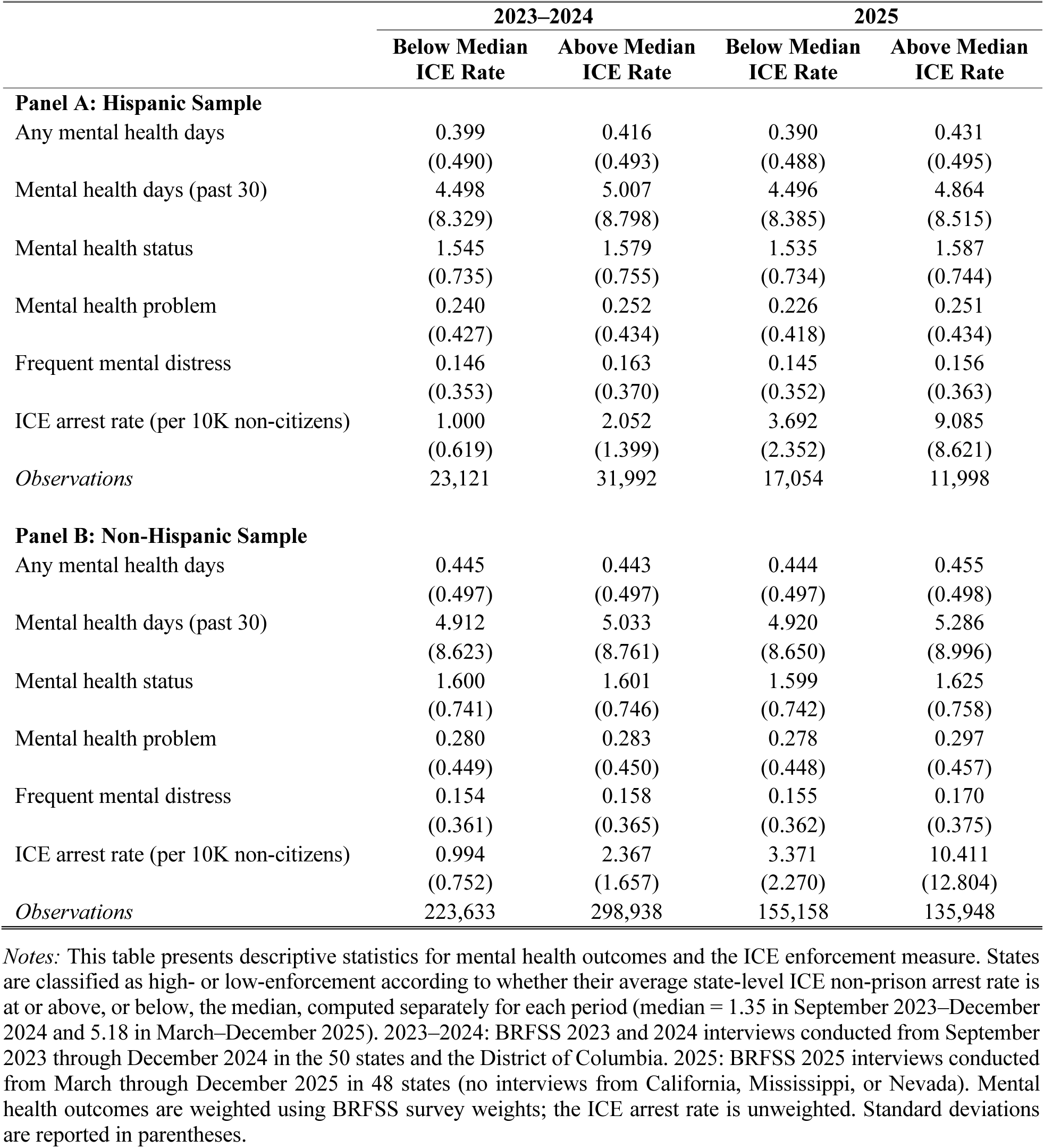
Descriptive Statistics by ICE Enforcement Intensity.

|  | 2023–2024 |  | 2025 |  |
|---|---|---|---|---|
|  | **Below Median ICE Rate** | **Above Median ICE Rate** | **Below Median ICE Rate** | **Above Median ICE Rate** |
| **Panel A: Hispanic Sample** |  |  |  |  |
| Any mental health days | 0.399 | 0.416 | 0.390 | 0.431 |
|  | (0.490) | (0.493) | (0.488) | (0.495) |
| Mental health days (past 30) | 4.498 | 5.007 | 4.496 | 4.864 |
|  | (8.329) | (8.798) | (8.385) | (8.515) |
| Mental health status | 1.545 | 1.579 | 1.535 | 1.587 |
|  | (0.735) | (0.755) | (0.734) | (0.744) |
| Mental health problem | 0.240 | 0.252 | 0.226 | 0.251 |
|  | (0.427) | (0.434) | (0.418) | (0.434) |
| Frequent mental distress | 0.146 | 0.163 | 0.145 | 0.156 |
|  | (0.353) | (0.370) | (0.352) | (0.363) |
| ICE arrest rate (per 10K non-citizens) | 1.000 | 2.052 | 3.692 | 9.085 |
|  | (0.619) | (1.399) | (2.352) | (8.621) |
| *Observations* | 23,121 | 31,992 | 17,054 | 11,998 |
| **Panel B: Non-Hispanic Sample** |  |  |  |  |
| Any mental health days | 0.445 | 0.443 | 0.444 | 0.455 |
|  | (0.497) | (0.497) | (0.497) | (0.498) |
| Mental health days (past 30) | 4.912 | 5.033 | 4.920 | 5.286 |
|  | (8.623) | (8.761) | (8.650) | (8.996) |
| Mental health status | 1.600 | 1.601 | 1.599 | 1.625 |
|  | (0.741) | (0.746) | (0.742) | (0.758) |
| Mental health problem | 0.280 | 0.283 | 0.278 | 0.297 |
|  | (0.449) | (0.450) | (0.448) | (0.457) |
| Frequent mental distress | 0.154 | 0.158 | 0.155 | 0.170 |
|  | (0.361) | (0.365) | (0.362) | (0.375) |
| ICE arrest rate (per 10K non-citizens) | 0.994 | 2.367 | 3.371 | 10.411 |
|  | (0.752) | (1.657) | (2.270) | (12.804) |
| *Observations* | 223,633 | 298,938 | 155,158 | 135,948 |

*Notes:* This table presents descriptive statistics for mental health outcomes and the ICE enforcement measure. States are classified as high- or low-enforcement according to whether their average state-level ICE non-prison arrest rate is at or above, or below, the median, computed separately for each period (median = 1.35 in September 2023–December 2024 and 5.18 in March–December 2025). 2023–2024: BRFSS 2023 and 2024 interviews conducted from September 2023 through December 2024 in the 50 states and the District of Columbia. 2025: BRFSS 2025 interviews conducted from March through December 2025 in 48 states (no interviews from California, Mississippi, or Nevada). Mental health outcomes are weighted using BRFSS survey weights; the ICE arrest rate is unweighted. Standard deviations are reported in parentheses.

Supplemental Table S1 reports sociodemographic characteristics. Compared to non-Hispanic adults, Hispanic adults reported lower educational attainment (58–60% vs. 34% with a high school education or less), lower incomes (58–59% vs. 34–35% with household income below $50,000), and lower rates of health insurance coverage (76% vs. 94-95%). State-level arrest rates rose sharply between the two periods. The median rate increased from 1.35 to 5.18 arrests per Figure 2 and Table 2 presents the main results for the period 2023-2024 from models regressing mental health outcomes (described in detail in Methods) against the rate of non-prison ICE arrests in the respondents’ state and month of interview, state and calendar month fixed effects, and individual covariates (estimating equation (1) in Methods) separately for Hispanic and non-Hispanic samples, respectively. In Panel A of Figure 2, higher immigration enforcement is associated with increases in poor mental health for Hispanic adults at both the extensive margin (reporting any poor mental health days or mental health problems) and the intensive margin (the number of poor mental health days). These estimates are statistically significant (except for frequent distress). By contrast, Panel B of Figure 2, shows that the estimated coefficients for the non-Hispanic sample are generally closer to zero with inconsistent signs; most estimates for this group are not statistically distinguishable from zero, suggesting little systematic association between ICE arrests and mental health outcomes among non-Hispanic adults.

**Figure 1.**
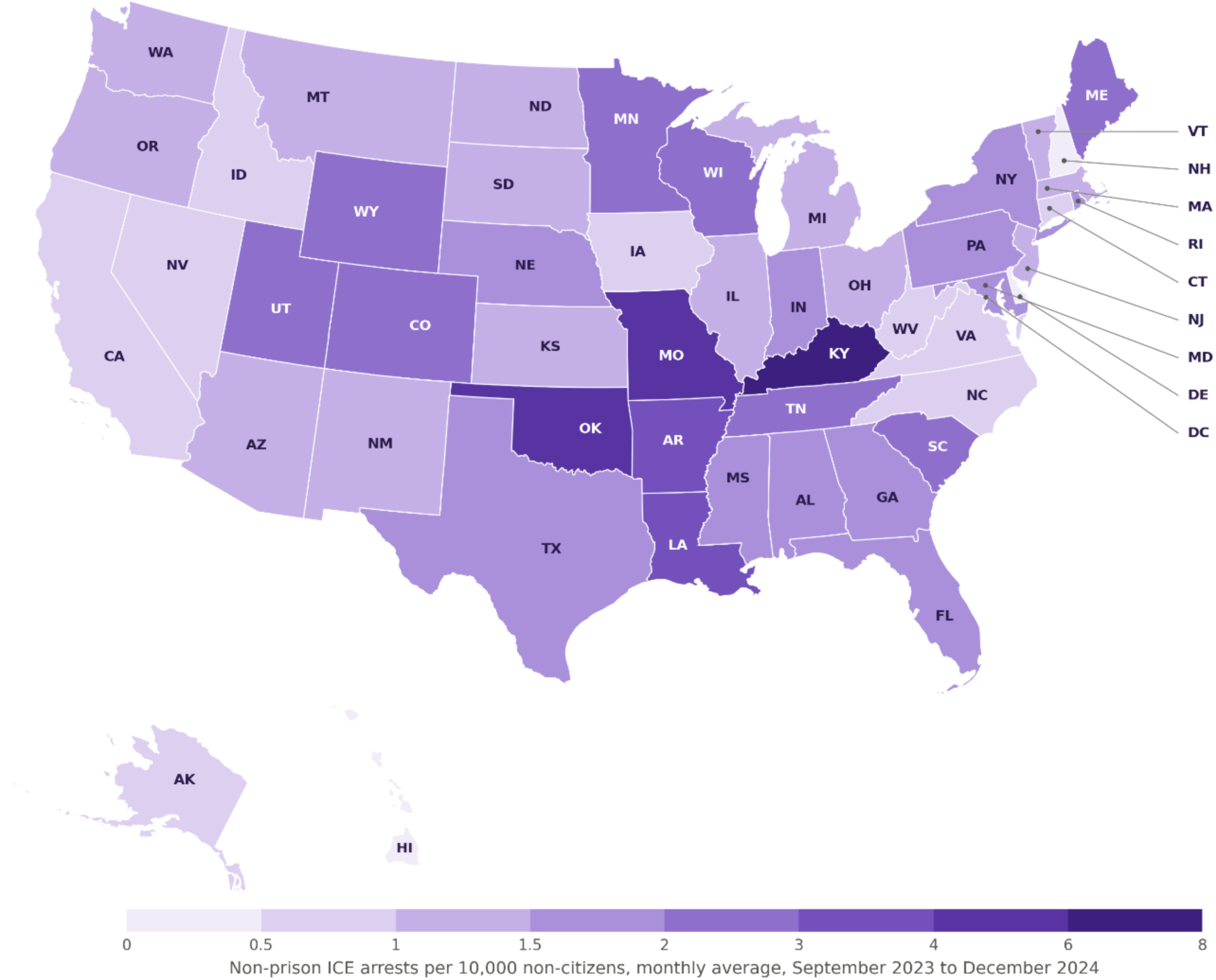
Non-Prison ICE Arrest Rate (per 10,000 Non-Citizens) *Notes:* This figure shows the average monthly non-prison ICE arrest rate per 10,000 non-citizens by state over the period September 2023 through December 2024. Darker shading indicates higher arrest rates.

**Figure 2.**
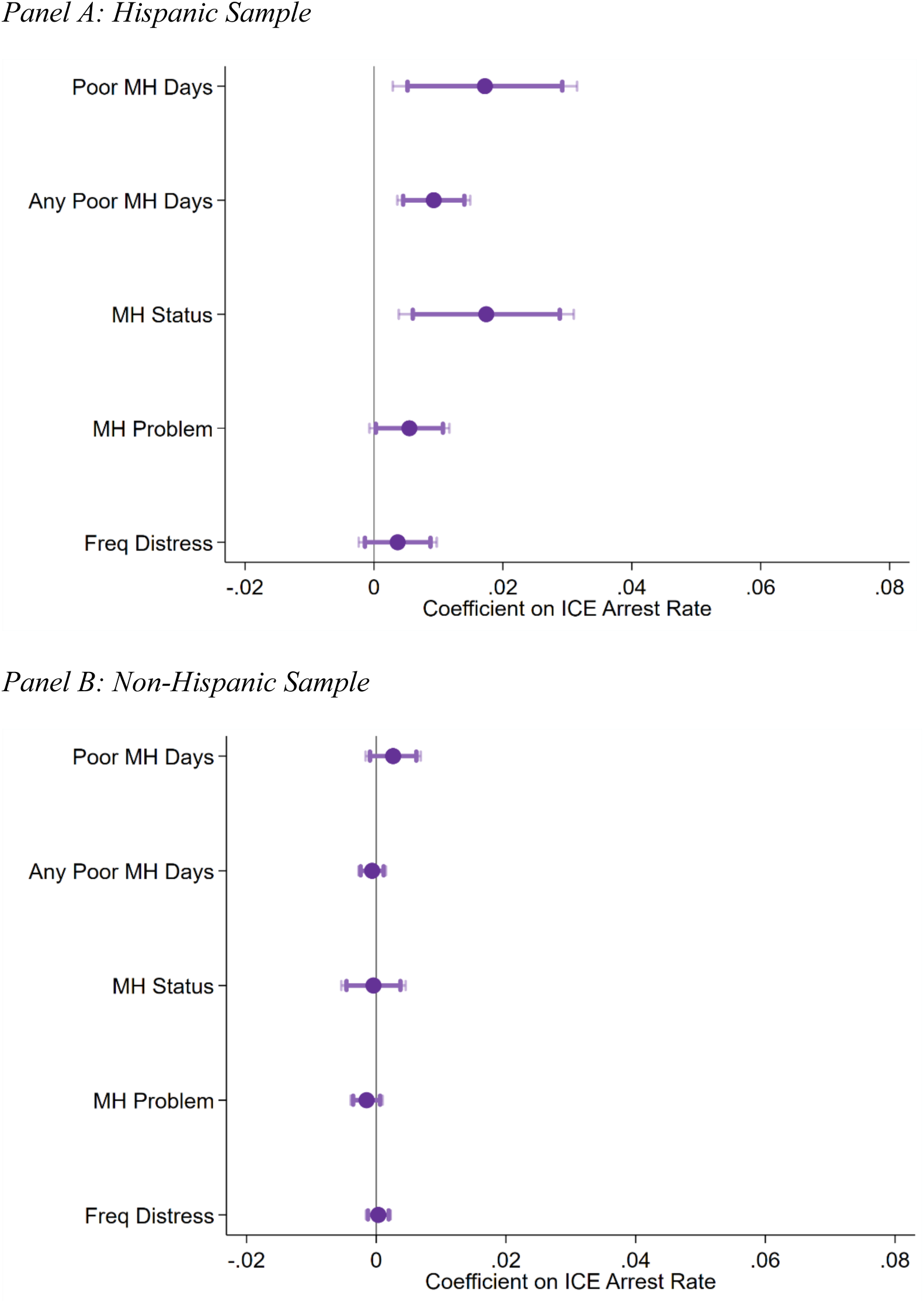
ICE Arrest Rates and Mental Health. *Notes:* This figure plots coefficient estimates of the association between ICE arrest rates and mental health outcomes among Hispanic and non-Hispanic respondents for the 2023-2024 sample. Dark and light shaded bars indicate 90% and 95% confidence intervals, respectively. All estimates use BRFSS survey weights and include sociodemographic controls (age, education, marital status, income categories, and health insurance) and the unemployment rate. Standard errors are clustered at the state level. Count and ordinal outcome measures (MH Days and MH Status) are standardized to mean 0 and standard deviation 1.

**Table 2.**
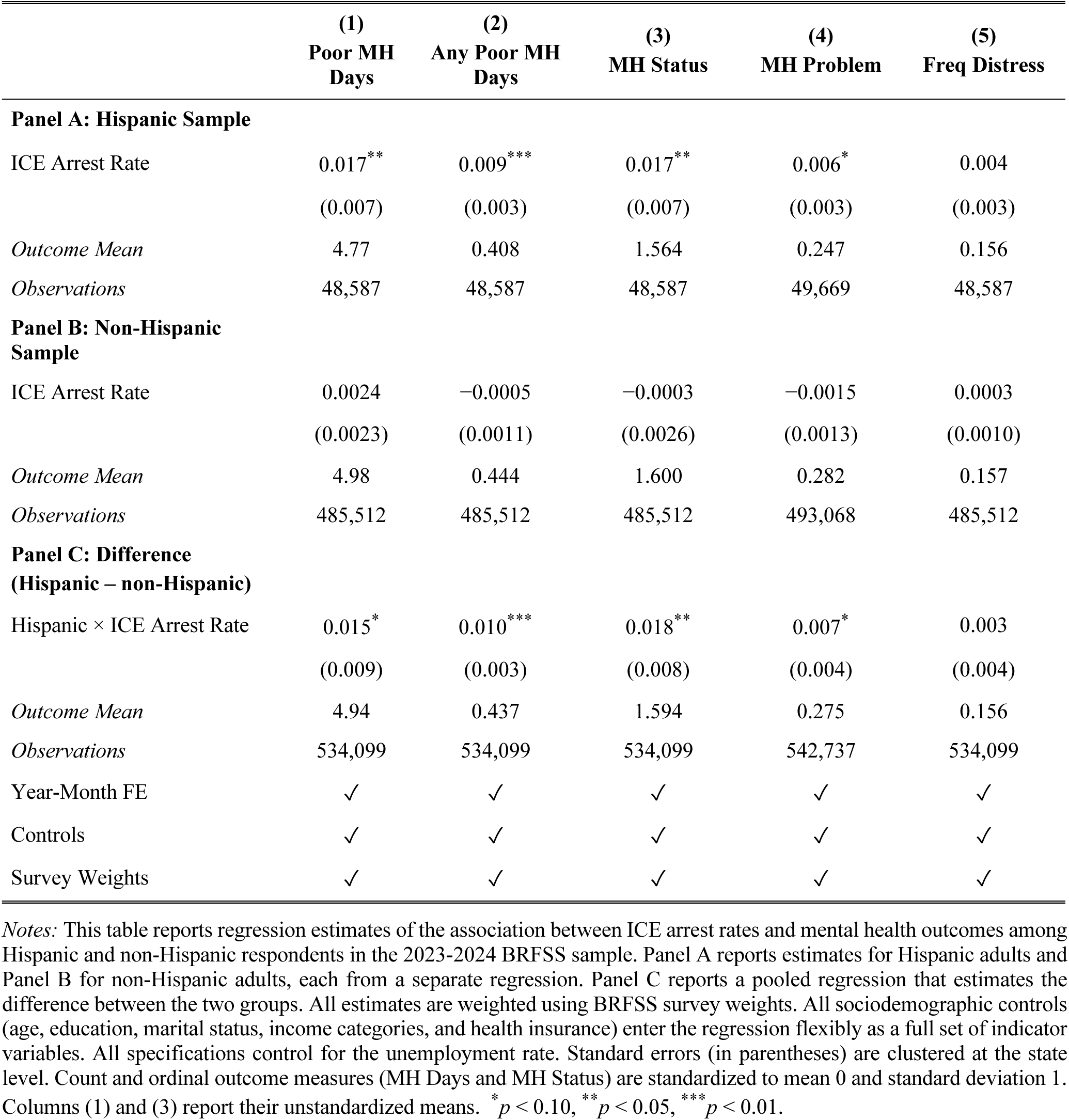
ICE Arrest Rates and Mental Health Among Hispanic and Non-Hispanic Adults.

|  | (1) | (2) | (3) | (4) | (5) |
|---|---|---|---|---|---|
|  | **Poor MH Days** | **Any Poor MH Days** | **MH Status** | **MH Problem** | **Freq Distress** |
|---|---|---|---|---|---|
| **Panel A: Hispanic Sample** |  |  |  |  |  |
| ICE Arrest Rate | 0.017<sup>**</sup> | 0.009<sup>***</sup> | 0.017<sup>**</sup> | 0.006<sup>*</sup> | 0.004 |
|  | (0.007) | (0.003) | (0.007) | (0.003) | (0.003) |
| *Outcome Mean* | 4.77 | 0.408 | 1.564 | 0.247 | 0.156 |
| *Observations* | 48,587 | 48,587 | 48,587 | 49,669 | 48,587 |
| **Panel B: Non-Hispanic Sample** |  |  |  |  |  |
| ICE Arrest Rate | 0.0024 | −0.0005 | −0.0003 | −0.0015 | 0.0003 |
|  | (0.0023) | (0.0011) | (0.0026) | (0.0013) | (0.0010) |
| *Outcome Mean* | 4.98 | 0.444 | 1.600 | 0.282 | 0.157 |
| *Observations* | 485,512 | 485,512 | 485,512 | 493,068 | 485,512 |
| **Panel C: Difference<br>(Hispanic – non-Hispanic)** |  |  |  |  |  |
| Hispanic × ICE Arrest Rate | 0.015<sup>*</sup> | 0.010<sup>***</sup> | 0.018<sup>**</sup> | 0.007<sup>*</sup> | 0.003 |
|  | (0.009) | (0.003) | (0.008) | (0.004) | (0.004) |
| *Outcome Mean* | 4.94 | 0.437 | 1.594 | 0.275 | 0.156 |
| *Observations* | 534,099 | 534,099 | 534,099 | 542,737 | 534,099 |
| Year-Month FE | ✓ | ✓ | ✓ | ✓ | ✓ |
| Controls | ✓ | ✓ | ✓ | ✓ | ✓ |
| Survey Weights | ✓ | ✓ | ✓ | ✓ | ✓ |

*Notes:* This table reports regression estimates of the association between ICE arrest rates and mental health outcomes among Hispanic and non-Hispanic respondents in the 2023-2024 BRFSS sample. Panel A reports estimates for Hispanic adults and Panel B for non-Hispanic adults, each from a separate regression. Panel C reports a pooled regression that estimates the difference between the two groups. All estimates are weighted using BRFSS survey weights. All sociodemographic controls (age, education, marital status, income categories, and health insurance) enter the regression flexibly as a full set of indicator variables. All specifications control for the unemployment rate. Standard errors (in parentheses) are clustered at the state level. Count and ordinal outcome measures (MH Days and MH Status) are standardized to mean 0 and standard deviation 1. Columns (1) and (3) report their unstandardized means. <sup>\*</sup>*p* < 0.10, <sup>\*\*</sup>*p* < 0.05, <sup>\*\*\*</sup>*p* < 0.01.

To assess the magnitude of these relationships, we turn to Table 2. Beginning with the primary outcome in Panel A, column (1) shows that a one-unit increase in the ICE arrest rate (corresponding to one additional non-prison arrest per 10,000 non-citizens) is associated with a 0.017 standard deviation increase in the number of poor mental health days reported in the past month (*p* < 0.05). The estimate in column (3) is comparable for mental health status, indicating a shift toward greater mental distress. To put this estimate in perspective, the standard deviation of poor mental health days in the sample is 8.6. Thus, a one-unit increase in the ICE arrest rate corresponds to approximately 0.15 additional poor mental health days per month (which is similar in magnitude to Bor et al. (2018), who found that each additional police killing was associated with 0.14 additional poor mental health days among Black Americans^20^). Because enforcement intensity varies meaningfully across states, a change in the ICE rate equal to the interquartile range (approximately 1.3) implies roughly 0.19 additional poor mental health days per month among Hispanic adults.

Column (2) of Table 2, Panel A reports results for the binary outcome of having any poor mental health days. The estimates suggest that a one-unit increase in the ICE arrest rate is associated with a 0.9 percentage point increase in the likelihood of reporting any poor mental health days (*p* < 0.01), or roughly a 2.2 percent increase relative to the outcome mean (0.408). In Column (3), we find a 0.017 standard-deviation increase in the mental health status index (*p* < 0.05), indicating a deterioration in mental health. Column (4) reports results for the composite mental health problem indicator, which combines a prior diagnosis of depressive disorder with frequent mental distress. We find that a one-unit increase in the ICE arrest rate is associated with a 0.6 percentage point increase in the likelihood of reporting a mental health problem (*p* < 0.10). Given that the mean of this outcome is 0.247, this corresponds to approximately a 2.4 percent increase relative to the outcome mean. Column (5) suggests a positive association between the ICE arrest rate and frequent mental distress; however, the estimate is not statistically significant.

In Table 2, Panel B, the estimated relationship between the ICE arrest rate and mental health outcomes for non-Hispanic adults is close to zero and statistically insignificant across all specifications. For example, in column (1), the coefficient is 0.0024 (*s.e.* = 0.0023), implying that we can rule out effects larger than roughly 0.007 standard deviations in absolute value at the 95 percent confidence level. The 95% confidence interval is 0.0024 ± 1.96 × 0.0023 = [−0.0021, 0.0069], implying that we can rule out associations larger than approximately 0.007 standard deviations in absolute value. This is an order of magnitude smaller than the corresponding estimate for Hispanic adults. Similarly small and statistically insignificant estimates appear in columns (2) through (5), with confidence intervals tightly centered around zero. These results suggest that the null findings for non-Hispanic adults are not driven by imprecision but rather reflect economically small and precisely estimated effects.

Panel C of Table 2 reports estimates from a pooled regression that interacts the ICE arrest rate with an indicator for Hispanic adults. The interaction coefficient tests whether the association between arrest rates and mental health differs between Hispanic and non-Hispanic adults. Because the non-Hispanic estimates are close to zero, the difference is similar in magnitude to the Hispanic estimates and is statistically significant in columns (1) through (4).

There is evidence of heterogeneity in estimates by sex, educational attainment, and race (online supplemental Figures S1, S2 and S3, and S4, respectively). We find significant increases in poor mental health among Hispanic women, but not among Hispanic men. We find no evidence that ICE arrests are associated with poor mental health among non-Hispanic women or non-Hispanic men. The estimates for women are larger on every mental health measure. Relative to each group’s mean, e.g., an additional non-prison arrest per 10,000 non-citizens is associated with a 3.3 percent increase in the likelihood of reporting a poor mental health day among Hispanic women, compared with 0.4 percent among Hispanic men. The difference between women and men is also statistically significant, implying a 1.3 percentage point larger increase for women (online supplemental Figure S3, Panel A). Stratifying the sample by educational attainment, we find that poor mental health is concentrated among Hispanic adults with a high school degree or less. Relative to Hispanic adults with at least some college, an additional arrest is associated with a 2.3 percentage point larger increase in the likelihood of reporting any poor mental health day among this group (online supplemental Figure S3, Panel B). Beyond the increase in poor mental health days, we find a statistically significant increase in frequent distress in this group translated into days, the association was equivalent to about 0.15 additional poor mental health days per month among Hispanic women and 0.36 among those with a high school education or less. In subgroup analyses by race, point estimates are larger for non-Hispanic Asian respondents compared to Hispanic respondents, but null or oppositely signed for non-Hispanic Black, American Indian, and other racial groups; however, small sample sizes preclude firm conclusions (Figure S4)

Figure 3 plots estimates using an event study approach that compares the evolution of mental health outcomes of residents before and after the month when their state experiences a positive month-to-month increase at or above the sample median compared to residents living in states that do not meet that criteria (see Methods). Pre-ICE activity estimates are small and statistically indistinguishable from zero for both Hispanic and non-Hispanic adults. A joint Wald test of the null that all pre-treatment coefficients equal zero fails to reject for either sample (Hispanic *p* = 0.352; non-Hispanic *p* = 0.250). Because a monotonic pre-treatment drift that continued into the post-treatment period could bias the estimates, we also fit a linear trend through the pre-treatment coefficients and test whether its slope is zero. The slope is statistically insignificant for both samples (Hispanic *p* = 0.964; non-Hispanic *p* = 0.339), and the point estimate for Hispanic adults is essentially zero. In the months following the enforcement increase, poor mental health among Hispanic adults rises and remains significantly elevated relative to non-Hispanic adults. Estimates stay close to zero throughout for the non-Hispanic sample.

**Figure 3.**
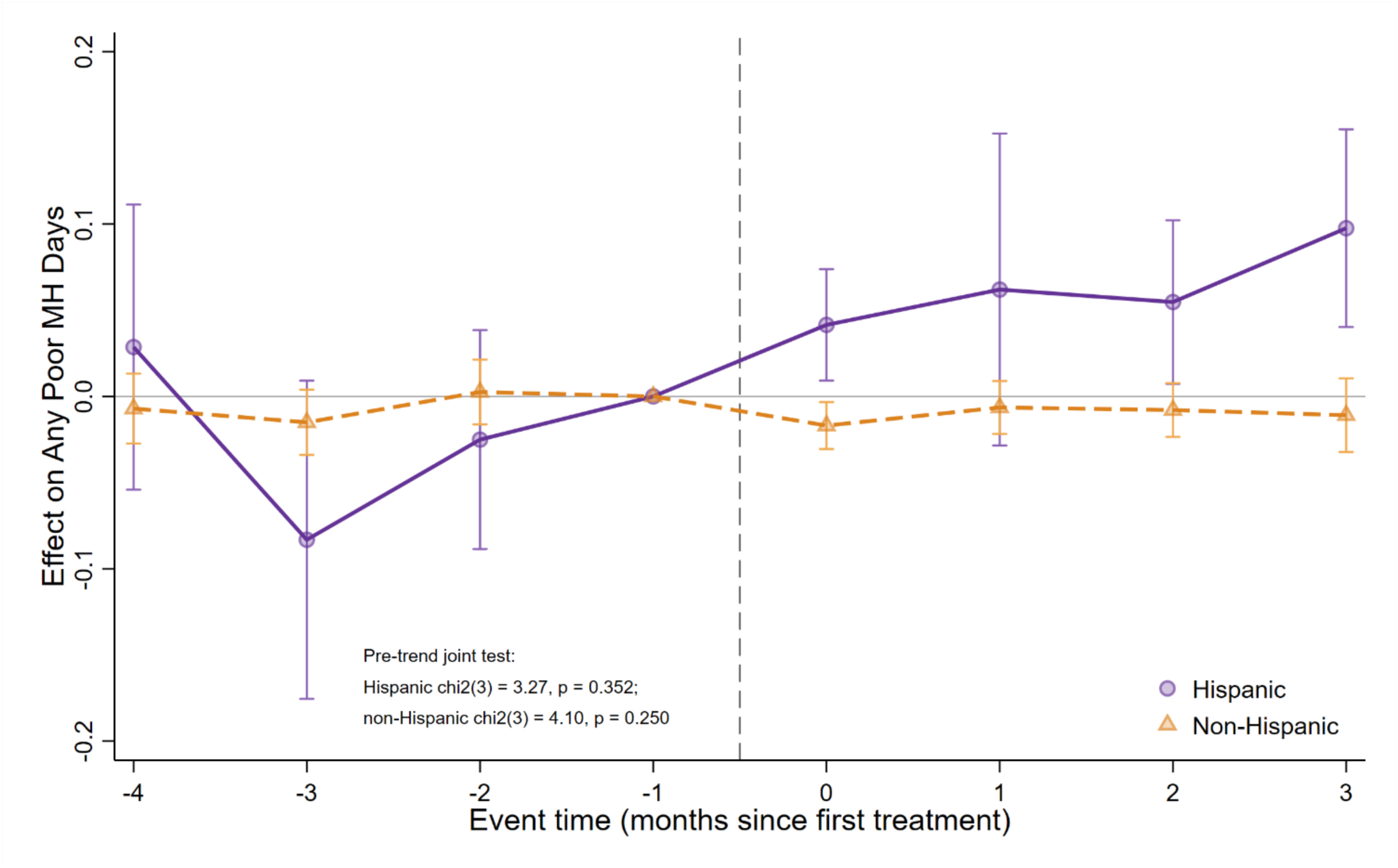
Event Study Estimates of the Effect of Large Increases in ICE Arrest Rates on Any Poor Mental Health Days by Hispanic Ethnicity. *Notes:* This figure presents heterogeneity-robust event study estimates of the association between large increases in state-level ICE arrest rates on the probability of reporting any poor mental health days in the past 30 days using the 2023-2024 BRFSS sample. Estimates are obtained using the augmented inverse-probability weighting estimator implemented via Stata’s `hdidregress` command on repeated cross-sectional data from the BRFSS. A state is classified as treated in the first month its ICE arrest rate experiences a positive month-to-month increase at or above the sample median, and is compared with states not-yet-treated in the same period. Models are estimated separately for Hispanic (purple circles) and non-Hispanic (orange triangles) adults. All models incorporate BRFSS survey weights and adjust for sex, age, education, marital status, household income, health insurance status, and the state unemployment rate. Standard errors are clustered at the state level. The month immediately before treatment serves as the reference period (normalized to zero). The figure reports a joint Wald test of the null that all pre-treatment coefficients equal zero. Vertical bars represent 95% confidence intervals.

Results from sensitivity analyses support the core findings. Lagged arrest rates and the two-month average of current and preceding month enforcement remain positively associated with worse mental health among Hispanic adults (online supplemental Table S2). The estimates also hold in models in which data from one state is omitted at a time (online supplemental Figure S5).

We then turn to the 2025 period, in which ICE arrests that are visible to the public rose substantially relative to prior years.^26,30^ We first analyzed larger contextual features of this period. Figure 4, Panel A displays the average co-movement of a state’s daily non-prison arrests with those of every other state, showing that arrests became increasingly nationally synchronized rather than locally driven. Average co-movement rises from 0.06 in 2023-2024 to 0.17 in 2025, and the share of state pairs moving together more than trivially rises from 4 percent to 35 percent. Figure 4, Panel B explores whether average mental health among BRFSS respondents differed between 2025 and 2023-2024 (Equation 2). Poor mental health increased in 2025 (any poor mental health days, 1.0 percentage points, *p* = 0.004), and the increase was similar across terciles of the change in state enforcement (1.1, 1.1, and 0.8 percentage points from the bottom to the top tercile).

**Figure 4.**
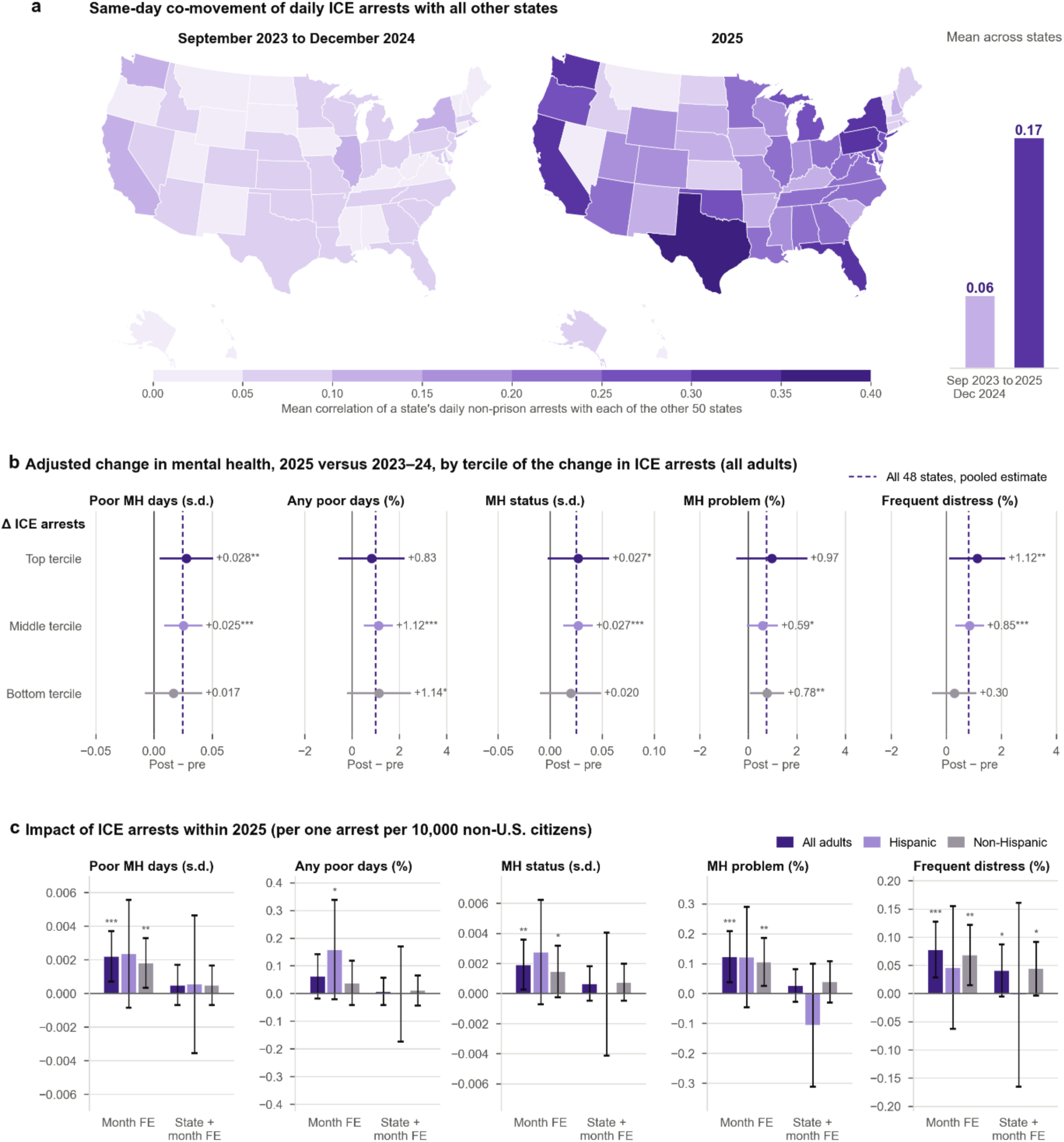
Non-Prison ICE Arrests and Mental Health in 2025. *Notes:* This figure presents three views of ICE enforcement and mental health in 2025 relative to 2023-2024. Panel A maps each state’s mean correlation of its daily non-prison ICE arrests with those of the other 50 states, after removing day-of-week means; darker shading indicates stronger same-day co-movement, and the bars report the mean across states. Panel B reports the regression-adjusted change in mental health between March-December 2023-24 and March-December 2025, estimated separately for the top, middle and bottom tercile of the change in a state’s monthly non-prison arrest rate per 10,000 non-citizens; the dashed line is the pooled estimate across all 48 states in a balanced panel. Panel C reports the coefficient on a state’s own monthly arrest rate within the March-December 2025 interviews, first with calendar-month fixed effects only and then adding state fixed effects. Panels B and C use BRFSS data with survey weights and control for demographics and the state unemployment rate. BRFSS 2025 does not include interviews from California, Mississippi, and Nevada. Continuous outcomes are standardized; binary outcomes are in percentage points. Bars and whiskers are 95% confidence intervals with standard errors clustered by state. \**p*<0.10, \*\**p*<0.05, \*\*\**p*<0.01.

Figure 4 Panel C then estimates the association between state-level ICE activity and mental health outcomes using variants of Equation (1). Conditioning on month fixed effects only (which allow for national variation in ICE arrests within a given month), the coefficient on ICE arrests is positive and statistically significant for four of the five outcomes. However, once state fixed effects are included – which restrict variation in ICE arrests to within state-month – the estimated coefficient is no longer distinguishable from zero.

## Discussion

We found that higher non-prison ICE arrest rates are associated with worse mental health outcomes among Hispanic – but not non-Hispanic – adults across multiple measures of mental distress. This relationship was concentrated among Hispanic adults with a high school diploma or less and among Hispanic women. Lower-educated individuals may face greater exposure to enforcement-related labor market disruptions, including job loss, reduced hours, or shifts into informal work, while having fewer financial buffers to absorb such shocks.^26^ Because financial insecurity is closely linked to anxiety and depressive symptoms, enforcement intensity may affect mental health not only through fear of detention but also through economic strain.^31^ The concentration of effects among women is consistent with enforcement generating stress through family and caregiving channels, as women disproportionately bear responsibility for managing household disruptions related to detention risk.^32–34^ We do not interpret this as evidence that enforcement has no effect on men’s mental health. The coefficient for Hispanic men is also positive, and the absence of significance may partly reflect differential nonresponse among the men at highest enforcement risk.

Our findings suggest that the social costs of contemporary immigration enforcement may extend beyond direct enforcement targets and economic disruptions. While the focus of our study was mental health, we note that, the impacts of enforcement may go beyond these outcomes. The stress of enforcement may result in coping behaviors such as alcohol use and, together with resource constraints, make healthy diets harder and regular access to health care difficult maintain.^35,36^ These pathways carry the effects of enforcement into physical health across communities, although we do not test them directly.

We also found that the relationship between non-prison ICE arrests and mental health changed between 2023-2024 and 2025. 2025 marked a different period from 2023–2024 in terms of the relationship between immigration enforcement and mental health in the United States. Not only did the volume of visible ICE arrests rise substantially, but arrests also became increasingly synchronized across states, reflecting a shift from local to national enforcement. Over the same period, poor mental health worsened nationwide regardless of the intensity of enforcement in a given state. The shift toward high-visibility, community-based arrests, combined with widespread national coverage, may have generated psychological spillovers extending beyond Hispanic communities to non-Hispanic adults, including U.S.-born individuals and legal residents. Consistent with these features, localized variation in enforcement intensity in 2025 was no longer correlated with mental health among Hispanic adults.

The findings for 2025 have implications for research on the health consequences of immigration enforcement moving forward. When enforcement is nationally salient, research designs that compare places identify only the local component of exposure and can miss its total effect. This is a more general limitation of geography-based studies of immigration enforcement outside of settings where local exposure stays meaningful (such as labor market outcomes). In addition, prevailing national dialogue and regime changes, which may shift *expectations* of enforcement, may impact health outcomes separately of actual enforcement. In these cases, measuring expectations is critical for better understanding the full consequences of immigration policy on health and well-being.

This study has several limitations. First, the BRFSS does not directly identify immigration status, preventing analysis by legal status and potentially masking heterogeneity among noncitizens. Second, the survey may underrepresent undocumented immigrants and others most exposed to enforcement risk. Evidence from public programs and household surveys suggests that immigration concerns may also discourage participation.^37–39^ If heightened enforcement disproportionately reduces participation among Hispanic adults experiencing greater psychological distress, our estimates could be attenuated. However, the direction of selection bias is uncertain. Third, although the Deportation Data Project includes sub-state geographic information for a subset of records, coverage is incomplete and inconsistent. Combined with the fact that the public-use BRFSS reports only state of residence, our analysis is necessarily conducted at the state-month level. Because immigration arrests are geographically concentrated within states, state-level measures may imperfectly capture respondents’ exposure to enforcement. This measurement error could attenuate the associations we estimate, but spatial aggregation alone does not establish the direction of bias. Relatedly, because the non-citizen denominator is held fixed within each year, states with large seasonal migrant populations may have mismeasured enforcement rates. Year-month fixed effects absorb common seasonal patterns, so this would bias estimates only if a state’s idiosyncratic seasonal population swings coincided with both enforcement timing and mental health.

## Methods

### Ethics Statement

No protocol approval was required for this study given the use of publicly-available data.

### Data Sources

We linked individual-level data from the 2023, 2024, and 2025 BRFSS with monthly state-level ICE arrest records from the Deportation Data Project (DDP), supplemented by state-month unemployment rates from the Federal Reserve Economic Data (FRED) database.

The BRFSS, administered by the Centers for Disease Control and Prevention (CDC), is the largest continuously conducted health survey in the United States, collecting data on health behaviors, chronic conditions, and preventive service use among adults aged 18 years and older via monthly telephone interviews, administered in English and Spanish, in all 50 states and the District of Columbia. In our baseline analysis, we use data from 2023 and 2024, which include approximately 890,000 respondents prior to sample restrictions. A key advantage of the BRFSS is that it records each respondent’s month of interview, enabling linkage to contemporaneous enforcement activity.

The primary outcome is the number of days during the past 30 days when the respondent’s mental health was “not good.” This widely used CDC Healthy Days measure captures stress, depression, and emotional problems.^20^ From it, we derived a binary indicator for any poor mental health days and a frequent mental distress indicator (≥14 days).^21,22^ We also used a three-level categorical mental health status variable (0 days, 1–13 days, ≥14 days) and a composite mental health problem indicator combining a lifetime depressive disorder diagnosis with current frequent distress. Relative to the frequent distress indicator, this measure additionally captures respondents with a diagnosed clinical history who are not currently in severe distress, situating current distress within a population at elevated baseline risk. Count and ordinal measures were standardized to mean zero and unit standard deviation. Demographic controls include sex, age, education, marital status, income, and insurance coverage. Approximately 10 percent of the sample identifies as Hispanic, the primary group of interest.

The DDP provides anonymized ICE Enforcement and Removal Operations arrest records obtained through Freedom of Information Act requests, including the date, state, and method of apprehension. We distinguish arrests occurring in prisons/jails from those in community settings.

### Study Population

We cleaned and geocoded arrest records to construct a state-by-month panel of ICE arrests for September 2023 through December 2024 and for January through December 2025, the latter from the September 2026 release of the Data Deportation Project. Our 2025 analyses use arrests from March through December 2025, the months in which the 2025 BRFSS was in the field across states. Some arrest records lacked state identifiers; we recovered these following the geocoding procedure in Aslim et al. (2026), using ICE Area of Responsibility and landmark information.^26^ Although the DDP provides records dating back to October 2011, earlier observations lack sufficient geographic identifiers for state-level assignment. We focused on non-prison arrests, which are more visible to communities than custodial transfers from correctional facilities. Non-prison arrests included worksite raids, home enforcement operations, and arrests in public locations such as outside courthouses. Prison-based arrests occur within correctional facilities following sentencing and are less visible to the public.

We measured enforcement intensity as the number of non-prison arrests per 10,000 non-U.S. citizens in each state and month, using American Community Survey one-year estimates as the denominator. Put differently, the annual ACS estimate is applied uniformly across all months of the year, so the denominator is fixed within state-year. Because 2025 ACS estimates were not yet available when we conducted the analysis, we linearly extrapolated each state’s 2025 non-citizen population from its 2023 and 2024 estimates. The non-citizen population included both lawfully present non-citizens and unauthorized immigrants. Although ICE primarily targets unauthorized immigrants, the broader non-citizen population provides a practical denominator because enforcement-related fear may affect the entire non-citizen community and because unauthorized status cannot be directly observed in census data. The mean ICE arrest rate was 1.78 per 10,000 non-citizens, with substantial cross-state variation (Figure 1). This variation formed the basis of the identification strategy.

The 2023–2024 sample includes 577,684 adults interviewed between September 2023 and December 2024 in all 50 states and the District of Columbia. After item nonresponse, the estimation sample is 534,099 (48,587 Hispanic and 485,512 non-Hispanic adults). The 2025 sample includes 320,158 adults interviewed between March and December 2025 in 48 states (the 2025 BRFSS has no interviews from California, Mississippi, or Nevada), with an estimation sample of 295,244 (25,499 Hispanic and 269,745 non-Hispanic adults).

### Statistical Analysis

We linked individual-level data from the BRFSS to monthly state-level non-prison ICE arrest rates constructed from the Deportation Data Project. Because the BRFSS records the month of interview, we constructed a repeated cross-sectional dataset at the monthly level, allowing us to leverage within-state month-to-month variation in ICE activity while controlling for year-month fixed effects to account for seasonality in mental health outcomes. Each respondent was matched to the ICE arrest rate in their state and month of interview, measured as non-prison arrests per 10,000 non-U.S. citizens. We estimated linear regression models of the form:

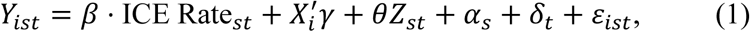

where *Y_ist_* is a mental health outcome for individual *i* in state *s* during year-month *t*. The specification included individual sociodemographic controls (*X_i_*: sex, age, education, marital status, income, and insurance coverage), the state-month unemployment rate (*Z_st_*), state fixed effects (*α_s_*), and year-month fixed effects (*δ_t_*).

The primary analysis focused on Hispanic adults given evidence that immigration enforcement disproportionately affects Hispanic communities through family, social, and community networks.^23,24^ We estimated the same models for non-Hispanic adults as a comparison group. We further examined heterogeneity by educational attainment and sex. We hypothesized that enforcement would disproportionately affect outcomes for women through caregiving and household channels: when household members face detention risk, women disproportionately bear the burden of managing family disruption, and enforcement may discourage utilization of health care, schools, and social services due to fear of exposure.^25^

We conducted several sensitivity analyses. First, we estimated a complementary difference-in-differences, event study specification,^27^ in which a state is classified as treated in the first month its ICE arrest rate experiences a positive month-to-month increase at or above the sample median. The specification compares differential changes in the outcome before and after treatment against the same changes in not-yet-treated states. This approach allows us to assess whether mental health was already changing prior to ICE activity as well as how mental health trajectories evolve after exposure. Because this analysis defines exposure differently from the main specification, comparing states around sharp increases in enforcement rather than using the continuous arrest rate, it complements rather than replaces our primary approach. Second, we estimated our primary models including lagged ICE arrest rates and leaving-one-out state at a time (to ensure that estimates were not driven by specific states where ICE activity may have been more salient). Additional details were provided in Appendix S1 in the Supplement.

We then conducted separate analyses using the 2025 BRFSS. We did so because 2025 marked the transition to a new Presidential administration (which previously has been linked to shifts in mental health independent of ICE activity) as well as more frequent and nationally-covered immigration enforcement.^28–30^ We posited that these conditions would make associations between state ICE activity and mental health either less salient, or harder to detect statistically. In addition, the 2025 BRFSS sample differed in that data collection started later (Spring 2025) and some states (California, Nevada, Mississippi – collectively comprising 27% of the Hispanic adult population) were not included.

We used three complementary analyses for 2025. First, to assess whether enforcement became nationally synchronized, we computed for each pair of states the correlation of daily non-prison arrest counts after removing each state’s day-of-week means, separately for September 2023– December 2024 and January–December 2025, and averaged each state’s correlations with the other 50 states. Second, to compare mental health across periods, we pooled BRFSS interviews conducted in March–December of 2023, 2024, and 2025 in the 48 states with interviews in both periods and estimated:

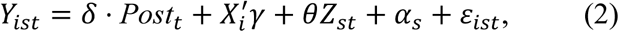

where *Post_t_* indicates an interview in 2025. We estimated Equation (2) for all 48 states and separately by tercile of the change in each state’s mean monthly non-prison arrest rate between the two periods. Third, within March–December 2025 interviews, we estimated Equation (1) with calendar-month fixed effects only and then with state fixed effects added.

All statistical analyses were conducted using Stata version 19 (StataCorp). The state map of arrest rates and the measure of cross-state synchronization in daily arrests were computed in Python 3.14 (pandas, NumPy, GeoPandas, Matplotlib). Standard errors were clustered at the state level. All analyses used BRFSS survey weights. All tests were two-sided; we report *p*-values throughout and define statistical significance as *p* < 0.05.

## Data Availability

All data and code for this study are available online at: https://dataverse.harvard.edu/dataset.xhtml?persistentId=doi:10.7910/DVN/P5PXZC

## Footnotes

1 The first category of each variable is the omitted reference category.

## Online Supplement

### Supplemental Appendix S1. Detailed Methods

To study the association between immigration enforcement and mental health, we estimate linear regression models that relate self-reported mental health outcomes to the intensity of ICE arrests in the respondent’s state and month of interview. The baseline specification takes the following form:

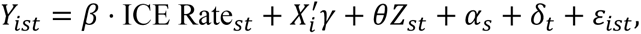

where *Y_ist_* is a mental health outcome (e.g., poor mental health days, mental health status, or frequent distress) for individual *i* residing in state *s* and interviewed in year-month *t*. The key explanatory variable, ICE Rate*_st_*, measures the number of non-prison ICE arrests per 10,000 non-U.S. citizens in state *s* during month *t*. The coefficient *β* captures the association between enforcement intensity and mental health outcomes. Beyond the primary mental health outcomes, we estimate the same models using measures of stress and general well-being to assess whether enforcement intensity is associated with a broader set of health-related outcomes.

The vector *X_i_* includes individual-level sociodemographic controls. To allow for flexibility in functional form, we include a full set of indicators for the following characteristics (including a missing category where applicable): sex (male, female), age (18–24, 25–29, 30–34, …, 80+), education (less than high school, high school graduate, some college, and college graduate), marital status (married, divorced, widowed, separated, never married, and unmarried couple), income (< $10,000, $10,000–$15,000, $15,000–$20,000, …, $200,000+), and health insurance coverage.^1^ To account for differences in economic conditions across states over time, we control for the state-month unemployment rate, denoted by *Z_st_*.

The specification includes state fixed effects (*α_s_*) to absorb time-invariant differences across states in mental health, demographics, immigration policy environments, and other unobserved factors. We also include year-month fixed effects (*δ_t_*) to control for national trends in mental health that are common across states, such as seasonal patterns in mental distress and nationwide changes in economic conditions or policy environments. Standard errors are clustered at the state level to account for serial correlation within states over time. All analyses use the BRFSS survey weights, which are the final respondent-level weights incorporating both landline and cellular telephone samples.

The primary analysis focuses on Hispanic adults, who represent approximately 10 percent of the sample. This focus is motivated by evidence that immigration enforcement disproportionately affects Hispanic communities, including U.S.-born individuals, through family, social, and community networks. Prior research also documents that immigration enforcement can generate fear, stress, and behavioral changes among Hispanic populations, even among those not directly targeted by enforcement actions. For these reasons, Hispanic respondents may be more exposed to the broader climate of immigration enforcement.

To assess whether associations between enforcement intensity and mental health are concentrated among populations more likely to be exposed to enforcement-related stress, we also estimate the same models separately for non-Hispanic adults. Comparing results across racial and ethnic groups provides a test of whether any observed associations are specific to communities more directly affected by immigration enforcement or instead reflect broader trends in mental health over time. We complement these analyses with heterogeneity-robust event-study specifications that align states by the first large increase in ICE arrest rates and compare treated states with states that are not-yet-treated in the same month. This approach allows us to examine whether mental health outcomes were evolving similarly across groups before treatment onset, providing an assessment of the parallel-trends assumption underlying the difference-in-differences design, and to characterize the timing and persistence of any post-treatment changes.

### Heterogeneity by Education and Sex

We examine heterogeneity by educational attainment because several channels could plausibly make the mental health consequences of enforcement intensity larger among lower-educated Hispanic adults. Lower-educated Hispanic adults are more likely to work in sectors characterized by informal employment, limited job protections, and greater exposure to workplace enforcement. Increased immigration enforcement may therefore heighten perceived job insecurity, reduce income stability, or disrupt household employment networks. Lower educational attainment is also associated with fewer financial buffers and more limited access to legal or informational resources, which could amplify stress responses to enforcement activity.

We also examine sex differences, estimating the models separately for men and women among Hispanic and non-Hispanic adults. Whether responses to immigration enforcement differ by sex is an empirical question, and existing work points in both directions. Women may experience enforcement-related stress differently because of caregiving responsibilities, household risk exposure, and differences in psychological responses to family and community instability. In turn, men may be more directly exposed to enforcement risk in labor markets and public spaces. Recent evidence also indicates that female-dominated sectors, such as childcare, may be affected by immigration enforcement, and that a broader chilling effect in the labor market may disproportionately affect immigrant women’s employment outcomes.^9^

Because the BRFSS does not include direct measures of these socioeconomics and household characteristics, we present them as potential channels rather than as empirically tested explanations.

**Supplemental Figure S1.**
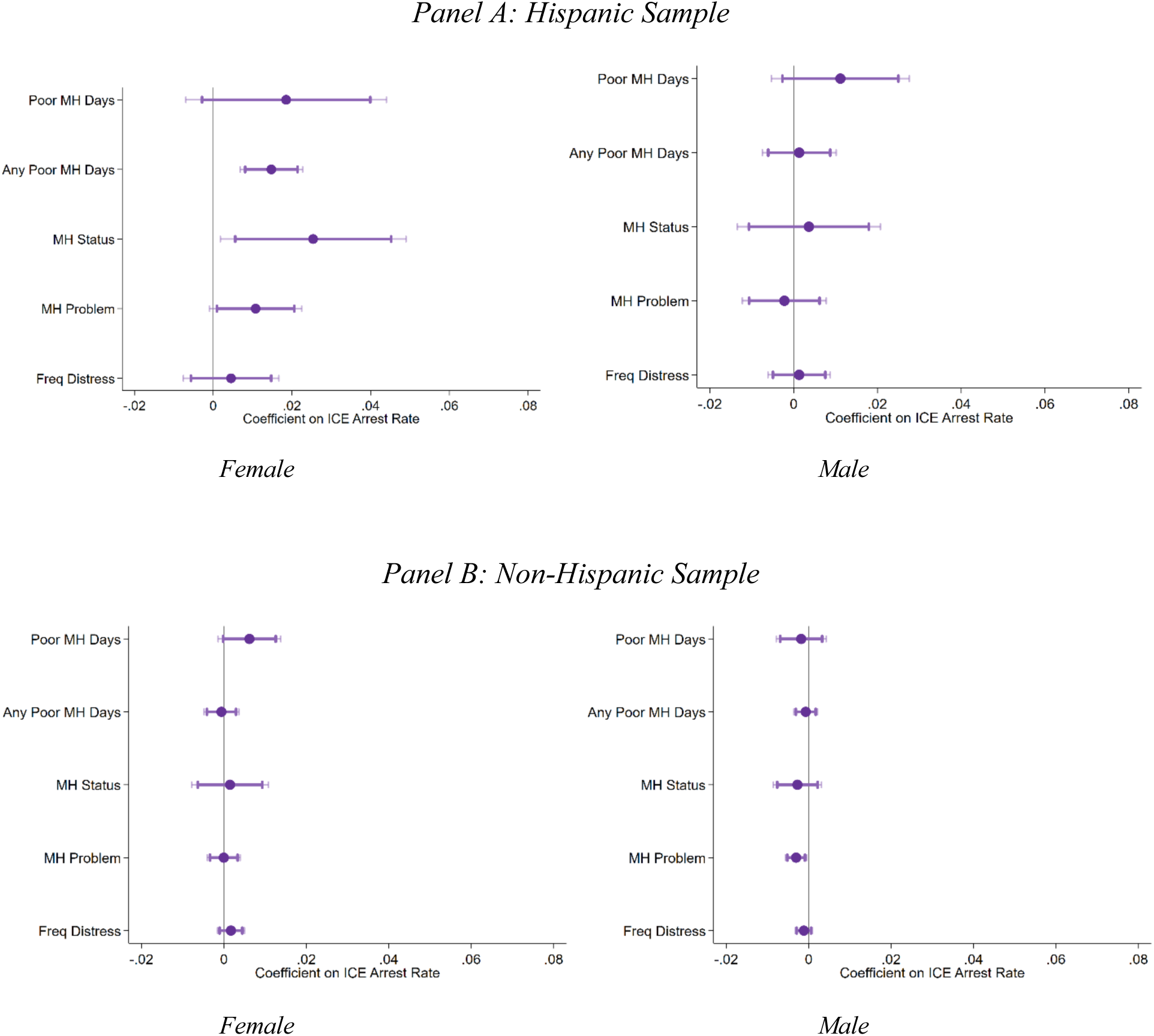
ICE Arrest Rates and Mental Health by Ethnicity and Sex. *Notes:* This figure plots coefficient estimates of the association between ICE arrest rates and mental health outcomes by ethnicity and sex in the 2023-2024 BRFSS sample. Dark and light shaded bars indicate 90% and 95% confidence intervals, respectively. All estimates use BRFSS survey weights and include sociodemographic controls (age, education, marital status, income categories, and health insurance) entered as indicator variables. Standard errors are clustered at the state level. Count and ordinal outcome measures (MH Days and MH Status) are standardized to mean 0 and standard deviation 1.

**Supplemental Figure S2.**
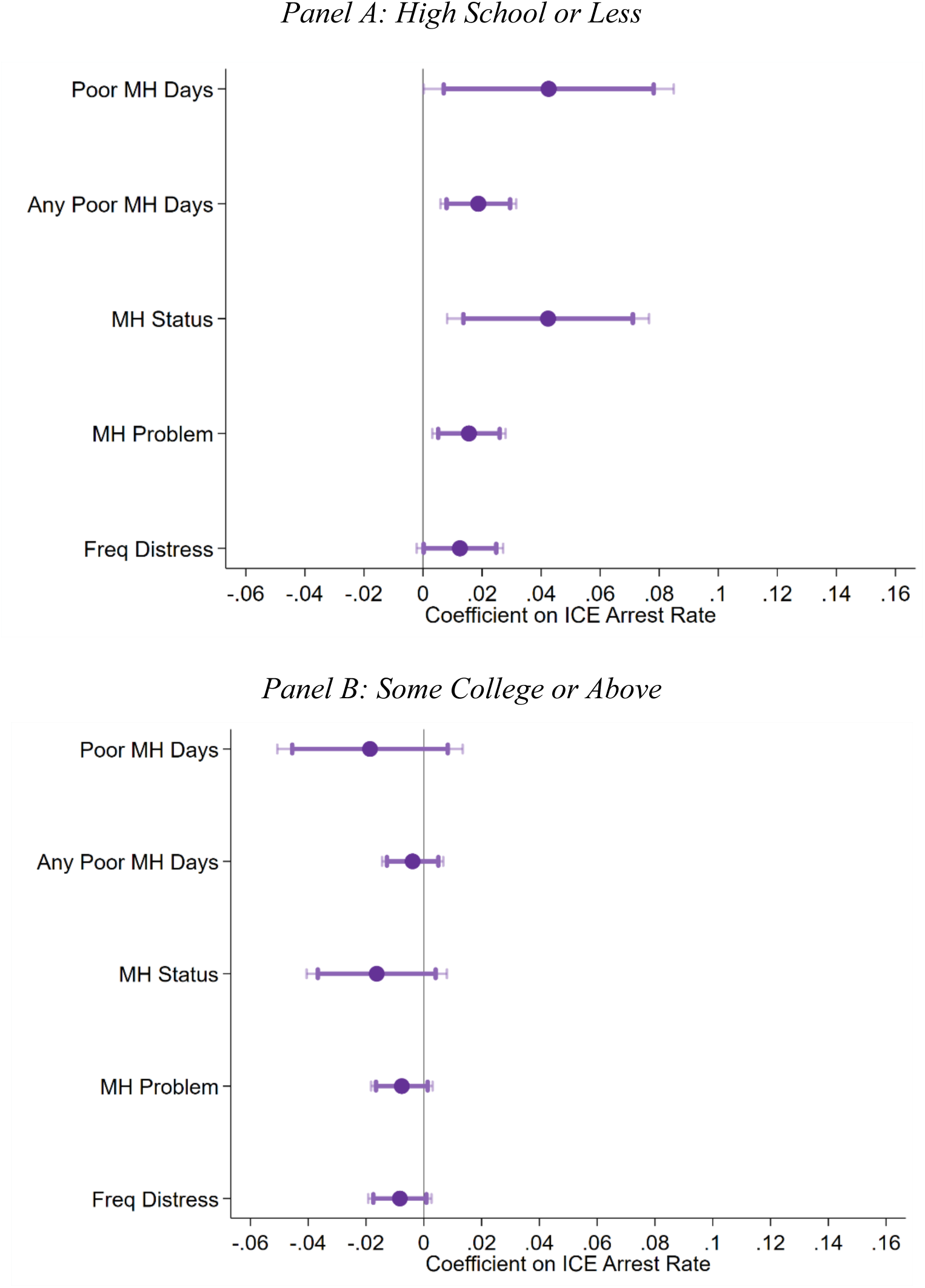
ICE Arrest Rates and Mental Health Among Hispanic Adults by Educational Attainment. *Notes:* This figure plots coefficient estimates of the association between ICE arrest rates and mental health outcomes among Hispanic adults by educational attainment in the 2023-2024 BRFSS sample. Low education is defined as high school diploma or less; high education is defined as some college or above. Dark and light shaded bars indicate 90% and 95% confidence intervals, respectively. All estimates use BRFSS survey weights and include sociodemographic controls (age, sex, marital status, income categories, and health insurance) entered as indicator variables. Standard errors are clustered at the state level. Count and ordinal outcome measures (MH Days and MH Status) are standardized to mean 0 and standard deviation 1.

**Supplemental Figure S3.**
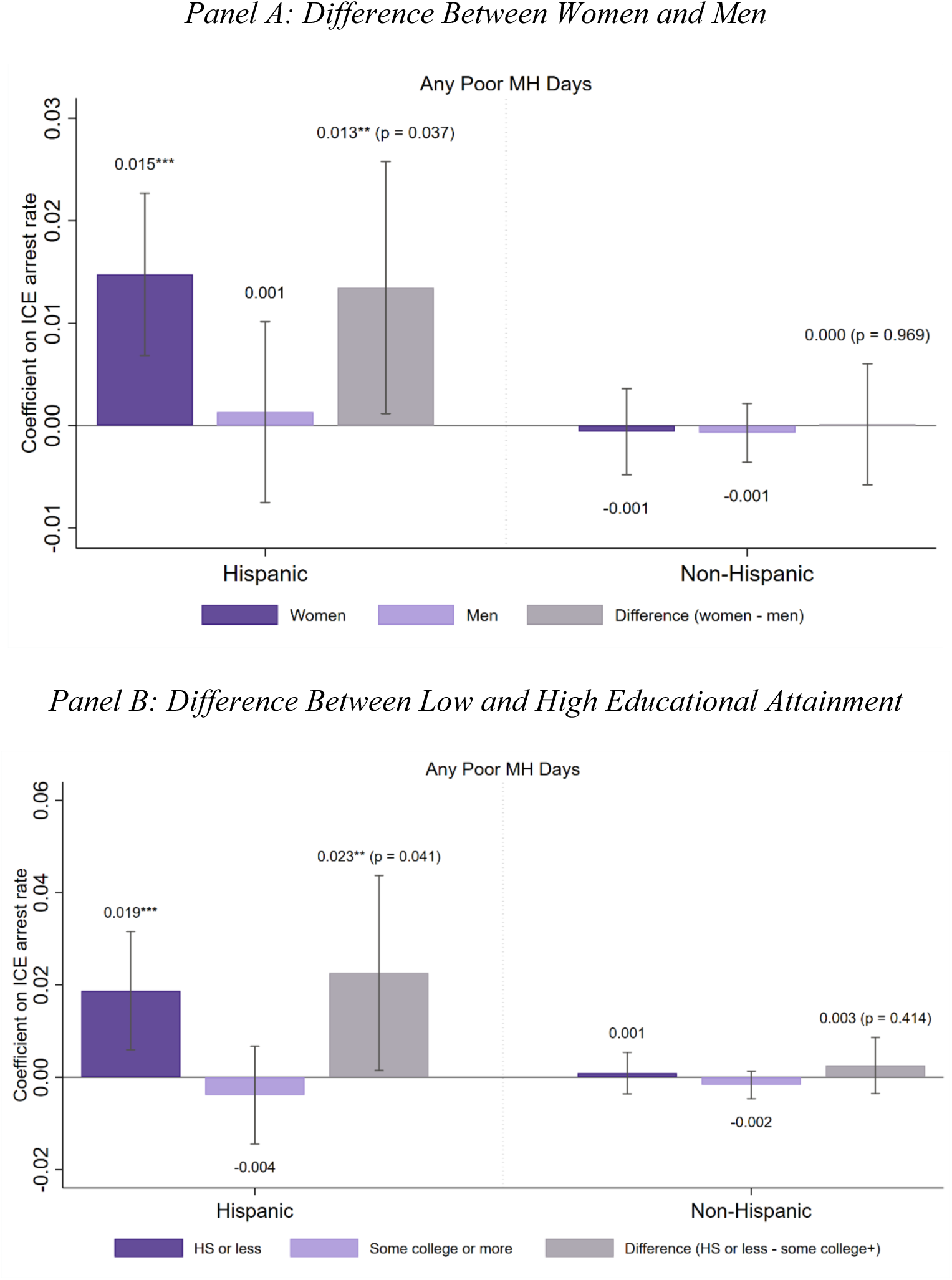
ICE Arrest Rates and Mental Health Among Hispanic Adults: Differences by Sex and Educational Attainment. *Notes:* This figure plots coefficient estimates of the association between ICE arrest rates and reporting any poor mental health day in the 2023-2024 BRFSS sample. We estimate separately by sex and educational attainment for Hispanic and non-Hispanic adults. The third bar in each panel is the difference between the two groups (women minus men in Panel A; high school or less minus some college or above in Panel B), and the *p*-value of this difference is reported above the bar. Whiskers indicate 95 percent confidence intervals. All estimates use BRFSS survey weights and include sociodemographic controls (age, sex, marital status, income categories, and health insurance) entered as indicator variables. Sex and educational attainment are excluded from the controls in the panel that defines groups by that characteristic. Standard errors are clustered at the state level. \**p* < 0.10, \*\**p* < 0.05, \*\*\**p* < 0.01.

**Supplemental Figure S4.**
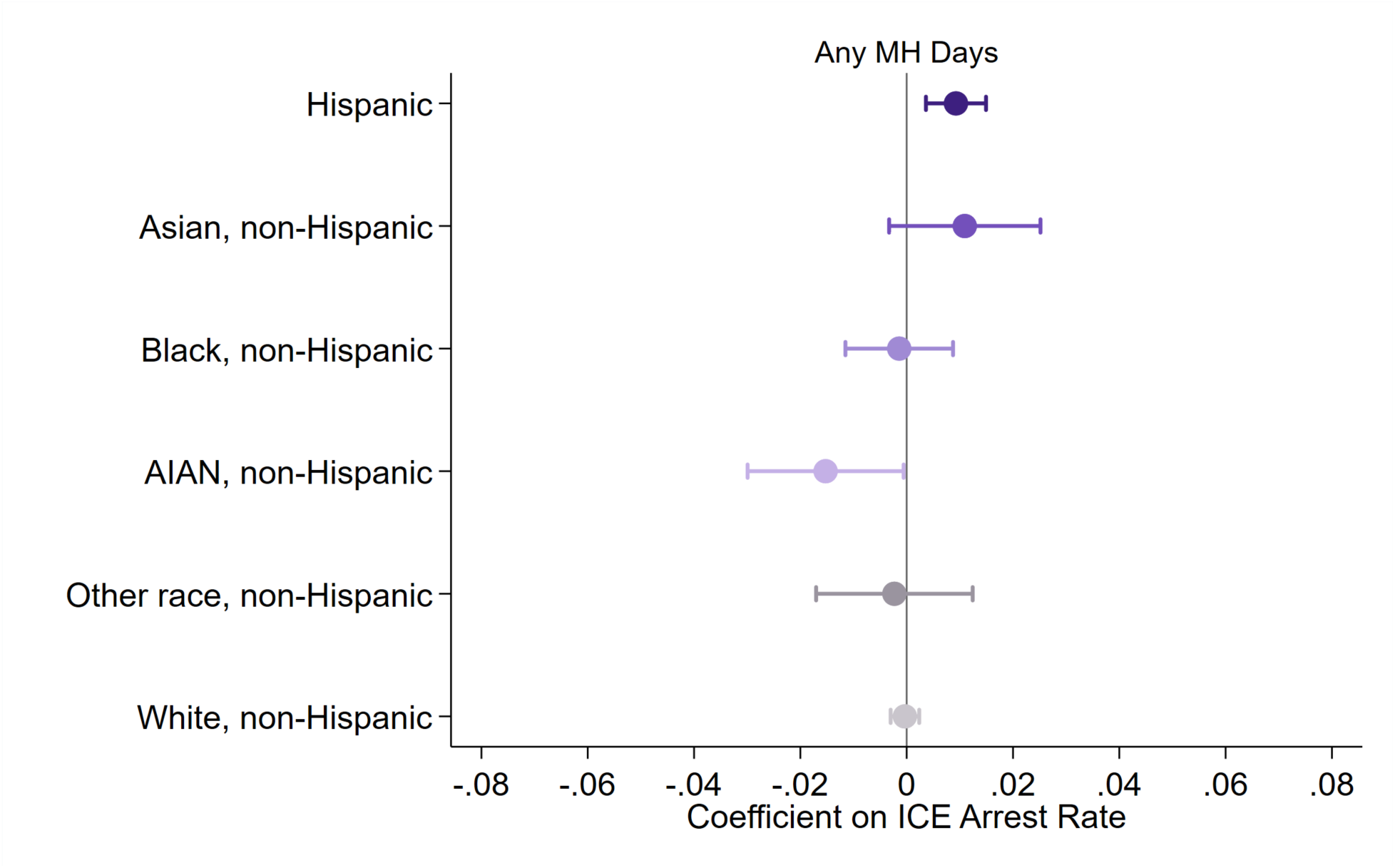
ICE Arrest Rates and Mental Health Across Racial and Ethnic Groups. *Notes:* This figure plots coefficient estimates of the association between ICE arrest rates and the outcome of any poor mental health days among Hispanic adults and across different non-Hispanic racial groups 2023-2024 BRFSS sample. Bars indicate 95% confidence intervals. All estimates use BRFSS survey weights and include sociodemographic controls (age, sex, marital status, income categories, and health insurance) entered as indicator variables. Standard errors are clustered at the state level.

**Supplemental Figure S5.**
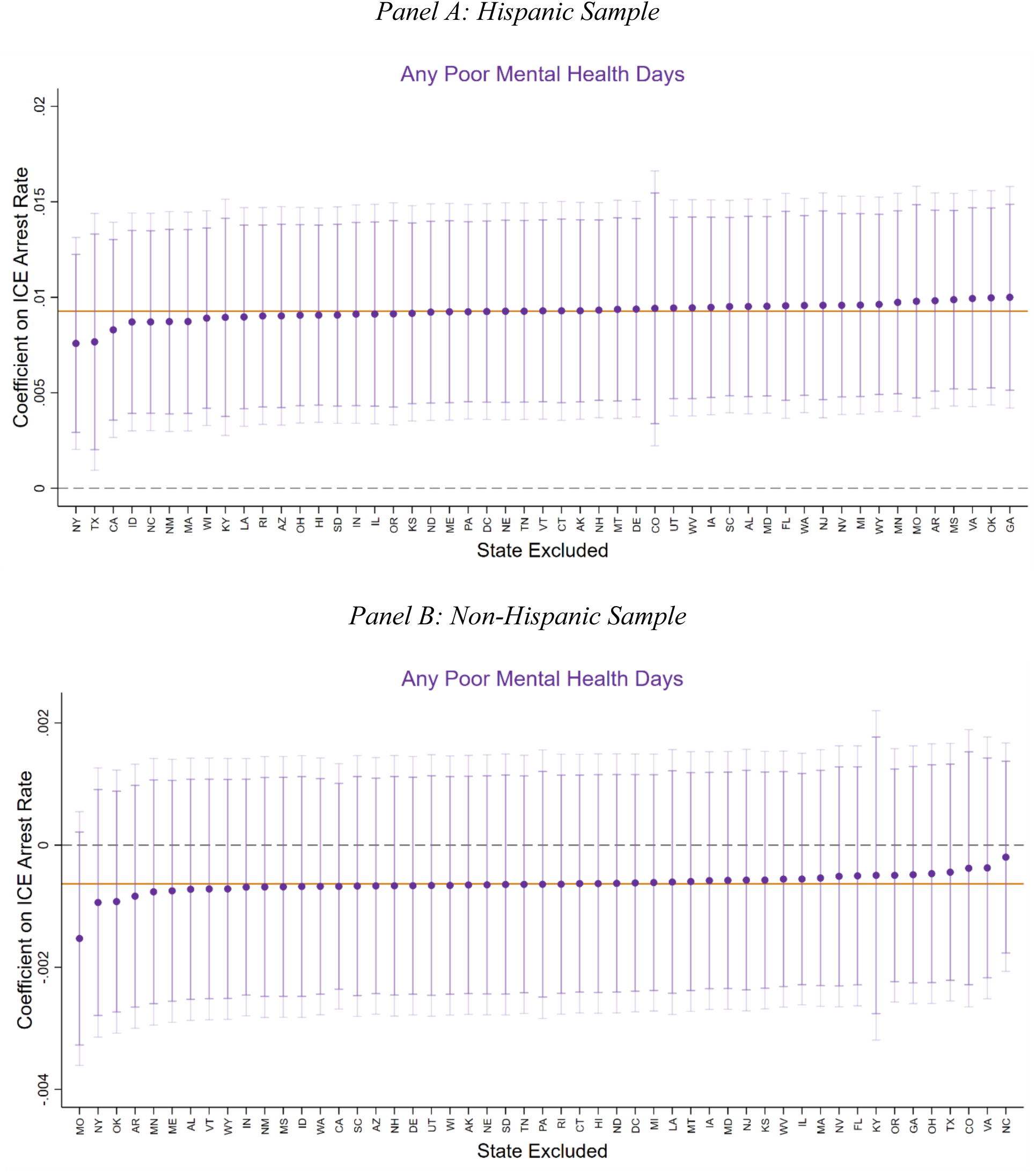
Leave-One-Out Analysis: Any Mental Health Days. *Notes:* This figure reports leave-one-out estimates of the association between ICE arrest rates and an indicator for any poor mental health days, excluding one state at a time, in the 2023-2024 BRFSS sample. The solid (orange) line denotes the baseline estimate reported in Table 2, and the dashed (gray) line indicates zero. Dark and light shaded bars indicate 90% and 95% confidence intervals, respectively.

**Supplemental Table S1.** Sociodemographic Characteristics.

|  | **2023–2024** |  | **2025** |  |
|---|---|---|---|---|
|  | **Hispanic** | **Non-Hispanic** | **Hispanic** | **Non-Hispanic** |
| Female | 0.504 | 0.511 | 0.509 | 0.511 |
|  | (0.500) | (0.500) | (0.500) | (0.500) |
| Married | 0.424 | 0.513 | 0.423 | 0.516 |
|  | (0.494) | (0.500) | (0.494) | (0.500) |
| Low education (HS or less) | 0.595 | 0.338 | 0.580 | 0.344 |
|  | (0.491) | (0.473) | (0.494) | (0.475) |
| Income < \$50K | 0.592 | 0.347 | 0.584 | 0.340 |
|  | (0.492) | (0.476) | (0.493) | (0.474) |
| Has health insurance | 0.762 | 0.947 | 0.764 | 0.941 |
|  | (0.426) | (0.224) | (0.424) | (0.235) |
| Unemployment rate | 3.895 | 3.626 | 4.095 | 3.924 |
|  | (0.734) | (0.774) | (0.647) | (0.743) |
| *Observations* | 55,113 | 522,571 | 29,052 | 291,106 |

*Notes:* This table reports descriptive statistics for sociodemographic characteristics of Hispanic and non-Hispanic respondents. 2023–2024: BRFSS 2023 and 2024 interviews conducted from September 2023 through December 2024 in the 50 states and the District of Columbia. 2025: BRFSS 2025 interviews conducted from March through December 2025 in 48 states (no interviews from California, Mississippi, or Nevada). Survey-level variables are weighted using BRFSS survey weights; the unemployment rate (state-month) is unweighted. Income < \$50K excludes respondents who did not know or refused to report income. Standard deviations are reported in parentheses. Observations are all respondents; item nonresponse varies across rows.

**Supplemental Table S2.** ICE Arrest Rates and Mental Health among Hispanic Adults: Lagged and Two-Month Average Exposure.

|  | **(1)**<br>**Poor MH Days** | **(2)**<br>**Any Poor MH Days** | **(3)**<br>**MH Status** | **(4)**<br>**MH Problem** | **(5)**<br>**Freq Distress** |
|---|---|---|---|---|---|
| **Panel A: Preceding month** |  |  |  |  |  |
| ICE Arrest Rate (*t* − *1*) | 0.021<sup>**</sup> | 0.008<sup>*</sup> | 0.018<sup>**</sup> | 0.010<sup>**</sup> | 0.006<sup>*</sup> |
|  | (0.009) | (0.004) | (0.008) | (0.004) | (0.003) |
| **Panel B: Two-month average** |  |  |  |  |  |
| Average ICE Arrest Rate (*t, t* − *1*) | 0.028<sup>**</sup> | 0.012<sup>**</sup> | 0.025<sup>**</sup> | 0.013<sup>**</sup> | 0.007 |
|  | (0.013) | (0.005) | (0.011) | (0.006) | (0.005) |
| *Observations* | 45,785 | 45,785 | 45,785 | 45,785 | 45,785 |
| State FE | ✓ | ✓ | ✓ | ✓ | ✓ |
| Year-Month FE | ✓ | ✓ | ✓ | ✓ | ✓ |
| Controls | ✓ | ✓ | ✓ | ✓ | ✓ |
| Survey Weights | ✓ | ✓ | ✓ | ✓ | ✓ |

*Notes:* This table reports regression estimates of the association between lagged ICE arrest rates and mental health outcomes among Hispanic respondents in the 2023-2024 BRFSS sample. Panel A uses enforcement in the preceding month (*t*−*1*). Panel B uses the average of enforcement in the current and preceding months (*t* and *t*−*1*). All estimates are weighted using BRFSS survey weights. Sociodemographic controls (age, education, marital status, income categories, and health insurance) are included flexibly as a full set of indicator variables. All specifications control for the unemployment rate. Standard errors (in parentheses) are clustered at the state level. Count and ordinal outcome measures (MH Days and MH Status) are standardized to mean 0 and standard deviation 1. <sup>\*</sup>*p* < 0.10, <sup>\*\*</sup>*p* < 0.05, <sup>\*\*\*</sup>*p* < 0.01.

## References

[1] American Immigration Council. Immigration detention expansion in trump’s second term, January 2026. URL https://www.americanimmigrationcouncil.org/. Accessed January 2026.

[2] Chloe N East, Caitlin Patler, and Elizabeth Cox. ICE Arrests across Trump’s First and Second Terms: Variation in Targeting, Method, and Geography, 2026. National Bureau of Economic Research, Working Paper 34794.

[3] Rita Rubin. US patients getting ICEd-out of health care. JAMA. 2026;335(10):835–837. doi:10.1001/jama.2026.1067.

[4] Saadi A, Patler C, Langer P. Duration in immigration detention and health harms. JAMA Network Open. 2025;8(1):e2456164.

[5] Joyner K, Burner E, Axeen S, Murugan S, English K, Schneberk TW. Migration-related trauma among asylum seekers exposed to the Migrant Protection Protocols. JAMA Network Open. 2026;9(1):e2550786.

[6] Offidani-Bertrand C. “It unleashed all the worries we tried to calm down”: the Trump administration’s impact on the mental health of immigrant communities. SSM-Mental Health. 2023;3:100207.

[7] Dadras O, Hazratzai MS. The silent trauma: US immigration policies and mental health. The Lancet Regional Health–Americas 2025;44.

[8] Giuntella O, Lonsky J. The effects of DACA on health insurance, access to care, and health outcomes. Journal of Health Economics. 2020;72:102320.

[9] Cha BS, Enriquez LE, Ro A. Beyond access: psychosocial barriers to undocumented students’ use of mental health services. Social Science & Medicine. 2019;233:193–200.

[10] Jill D McLeigh. How do immigration and customs enforcement (ICE) practices affect the mental health of children? American Journal of Orthopsychiatry, 80(1):96, 2010.

[11] Cindy Juby and Laura E Kaplan. Postville: The effects of an immigration raid. Families in Society, 92(2):147–153, 2011.

[12] Julia Shu-Huah Wang and Neeraj Kaushal. Health and mental health effects of local immigration enforcement. International Migration Review, 53(4):970–1001, 2019.

[13] Miguel Pinedo and Carmen R Valdez. Immigration enforcement policies and the mental health of US citizens: Findings from a comparative analysis. American Journal of Community Psychology, 66(1-2):119–129, 2020.

[14] Mark L Hatzenbuehler, Seth J Prins, Morgan Flake, Morgan Philbin, M Somjen Frazer, Daniel Hagen, and Jennifer Hirsch. Immigration policies and mental health morbidity among Latinos: A state-level analysis. Social Science & Medicine, 174: 169–178, 2017.

[15] Emilie Bruzelius and Aaron Baum. The mental health of Hispanic/Latino Americans following national immigration policy changes: United states, 2014–2018. American Journal of Public Health, 109(12):1786–1788, 2019.

[16] Courtney E. Boen, Nick Graetz, Atheendar S. Venkataramani, and Robin Ortiz. The bodily scars of legal violence: local immigration enforcement, state immigrant policy, and health inequality. *Social Forces*, soaf181, 2025.

[17] Romina Tome, Marcos A. Rangel, Christina M. Gibson-Davis, and Laura Bellows. Heightened immigration enforcement impacts US citizens’ birth outcomes: Evidence from early ICE interventions in North Carolina. PLoS ONE, 16(2):e0245020, 2021.

[18] Abigail S. Friedman and Atheendar S. Venkataramani. Chilling effects: US immigration enforcement and health care seeking among Hispanic adults: Study examines the effects of US immigration enforcement and health care seeking among Hispanic adults. Health Affairs, 40(7):1056–1065, 2021.

[19] Florencia Torche and Catherine Sirois. Restrictive immigration law and birth outcomes of immigrant women. American Journal of Epidemiology, 188(1):24–33, 2019.

[20] Jacob Bor, Atheendar S Venkataramani, David R Williams, and Alexander C Tsai. Police killings and their spillover effects on the mental health of black Americans: a population-based, quasi-experimental study. The Lancet, 392(10144):302–310, 2018.

[21] David G Moriarty, Mathew M Zack, and Rosemarie Kobau. The centers for disease control and prevention’s healthy days measures–population tracking of perceived physical and mental health over time. Health and Quality of Life Outcomes, 1(1):37, 2003.

[22] S Lane Slabaugh, Mona Shah, Matthew Zack, Laura Happe, Tristan Cordier, Eric Havens, Evan Davidson, Michael Miao, Todd Prewitt, and Haomiao Jia. Leveraging health-related quality of life in population health management: the case for healthy days. Population Health Management, 20(1):13–22, 2017.

[23] Krista M Perreira and Juan M Pedroza. Policies of exclusion: implications for the health of immigrants and their children. Annual Review of Public Health, 40(1): 147–166, 2019.

[24] Asad L Asad. Latinos’ deportation fears by citizenship and legal status, 2007 to 2018. Proceedings of the National Academy of Sciences, 117(16):8836–8844, 2020.

[25] Joanna Dreby. The burden of deportation on children in Mexican immigrant families. Journal of Marriage and Family, 74(4):829–845, 2012.

[26] Erkmen G. Aslim, Janet Currie, Chris M. Herbst, and Erdal Tekin. How rising ice activity influences the childcare workforce. Proceedings of the National Academy of Sciences, 123(21), 2026.

[27] Andrew Goodman-Bacon. Difference-in-differences with variation in treatment timing. Journal of Econometrics, 225(2):254–277, 2021.

[28] Christopher Ojeda. Mental health in times of democratic backsliding: Insights from Trump’s second term. Journal of Health Politics, Policy and Law, 2026:12461779, 2026.

[29] Brittany N. Morey, San Juanita García, Tanya Nieri, Tim A. Bruckner, and Bruce G. Link. Symbolic disempowerment and Donald Trump’s 2016 presidential election: Mental health responses among Latinx and white populations. Social Science & Medicine, 289:114417, 2021.

[30] Chloe N. East, Caitlin Patler, and Elizabeth Cox. ICE arrests, 2015–2026: Variation in targeting, method, and geography. Proceedings of the National Academy of Sciences, 123(31):e2618096123

[31] Sarah Elshahat, Tina Moffat, and K. Bruce Newbold. Understanding the healthy immigrant effect in the context of mental health challenges: A systematic critical review. Journal of Immigrant and Minority Health, 24(6):1564–1579, 2022.

[32] Geri-Ann Galanti. The Hispanic family and male-female relationships: An overview. Journal of Transcultural Nursing, 14(3):180–185, 2003.

[33] Yamilé Molina, Vida Henderson, India J Ornelas, John R Scheel, Sonia Bishop, Sarah L Doty, Donald L Patrick, Shirley AA Beresford, and Gloria D Coronado. Understanding complex roles of family for Latina health: evaluating family obligation stress. Family & Community Health, 42(4):254–260, 2019.

[34] Hortensia Amaro, Nancy Felipe Russo, and Julie Johnson. Family and work predictors of psychological well-being among Hispanic women professionals. Psychology of Women Quarterly, 11(4):505–521, 1987.

[35] Chris Guerra, Eric J. Connolly, and Danielle L. Boisvert. The immigrant experience and alcohol use: Heart rate as a source of risk and resilience. Prevention Science, 24(2):309–321, 2023.

[36] M.N. LeCroy, R. Suss, R.G. Russo, S. Sifuentes, J.M. Beasley, R.G. Barajas-Gonzalez, P. Chebli, V. Foster, S.C. Kwon, C. Trinh-Shevrin, and S.S. Yi. Looking across and within: Immigration as a unifying structural factor impacting cardiometabolic health and diet. Ethnicity & Disease, 33(2–3):130–139, 2023.

[37] Marcella Alsan and Crystal S. Yang. Fear and the safety net: Evidence from secure communities. Review of Economics and Statistics, 106(6):1427–1441, 2024.

[38] Tara Watson. Inside the refrigerator: Immigration enforcement and chilling effects in Medicaid participation. American Economic Journal: Economic Policy, 6(3):313–338, 2014.

[39] J. David Brown and Misty L. Heggeness. Citizenship question effects on household survey response. Journal of Policy Analysis and Management, 45(1):e70004, 2026.

## Online Supplement -References

[1] Krista M Perreira and Juan M Pedroza. Policies of exclusion: implications for the health of immigrants and their children. Annual Review of Public Health, 40(1): 147–166, 2019.

[2] Asad L Asad. Latinos’ deportation fears by citizenship and legal status, 2007 to 2018. Proceedings of the National Academy of Sciences, 117(16):8836–8844, 2020.

[3] Nicole L Novak, Arline T Geronimus, and Aresha M Martinez-Cardoso. Change in birth outcomes among infants born to Latina mothers after a major immigration raid. International Journal of Epidemiology, 46(3):839–849, 2017.

[4] Kathleen M Roche, Rebecca MB White, Sharon F Lambert, John Schulenberg, Esther J Calzada, Gabriel P Kuperminc, and Todd D Little. Association of family member detention or deportation with Latino or Latina adolescents’ later risks of suicidal ideation, alcohol use, and externalizing problems. JAMA Pediatrics, 174(5):478–486, 2020.

[5] Amy L Johnson, Christopher Levesque, Neil A Lewis Jr, and Asad L Asad. Deportation threat predicts Latino US citizens and noncitizens’ psychological distress, 2011 to 2018. Proceedings of the National Academy of Sciences, 121(9):e2306554121, 2024.

[6] Joanna Dreby. Everyday illegal: When policies undermine immigrant families. University of California Press, 2015.

[7] William Paul Simmons, Cecilia Menjívar, and Elizabeth Salerno Valdez. The gendered effects of local immigration enforcement: Latinas’ social isolation in Chicago, Houston, Los Angeles, and Phoenix. International Migration Review, 55(1): 108–134, 2021.

[8] Umair Ali, Jessica H Brown, and Chris M Herbst. Secure communities as immigration enforcement: How secure is the child care market? Journal of Public Economics, 233:105101, 2024.

[9] Erkmen G. Aslim, Janet Currie, Chris M. Herbst, and Erdal Tekin. How rising ice activity influences the childcare workforce. Proceedings of the National Academy of Sciences, 123(21), 2026.

[10] Cynthia Bansak, Sarah Pearlman, and Chad Sparber. The impact of secure communities on the labor market outcomes of immigrant women. Journal of Policy Analysis and Management, 44(3):917–942, 2025.

